# A unified data infrastructure to support large-scale rare disease research

**DOI:** 10.1101/2023.12.20.23299950

**Authors:** Lennart F. Johansson, Steve Laurie, Dylan Spalding, Spencer Gibson, David Ruvolo, Coline Thomas, Davide Piscia, Fernanda de Andrade, Gerieke Been, Marieke Bijlsma, Han Brunner, Sandi Cimerman, Farid Yavari Dizjikan, Kornelia Ellwanger, Marcos Fernandez, Mallory Freeberg, Gert-Jan van de Geijn, Roan Kanninga, Vatsalya Maddi, Mehdi Mehtarizadeh, Pieter Neerincx, Stephan Ossowski, Ana Rath, Dieuwke Roelofs-Prins, Marloes Stok-Benjamins, K. Joeri van der Velde, Colin Veal, Gerben van der Vries, Marc Wadsley, Gregory Warren, Birte Zurek, Thomas Keane, Holm Graessner, Solve-RD consortium, Sergi Beltran, Morris A. Swertz, Anthony J. Brookes

**Author notes:** Shared last authors (Morris A. Swertz, Anthony J. Brookes). Full lists of all authors available at the end of this document: Solve-RD consortium.

## Abstract

The Solve-RD project brings together clinicians, scientists, and patient representatives from 51 institutes spanning 15 countries to collaborate on genetically diagnosing (“solving”) rare diseases (RDs). The project aims to significantly increase the diagnostic success rate by co-analysing data from thousands of RD cases, including phenotypes, pedigrees, exome/genome sequencing and multi-omics data. Here we report on the data infrastructure devised and created to support this co-analysis. This infrastructure enables users to store, find, connect, and analyse data and metadata in a collaborative manner. Pseudonymised phenotypic and raw experimental data are submitted to the RD-Connect Genome-Phenome Analysis Platform and processed through standardised pipelines. Resulting files and novel produced omics data are sent to the European Genome-phenome Archive, which adds unique file identifiers and provides long-term storage and controlled access services. MOLGENIS “RD3” and Café Variome “Discovery Nexus” connect data and metadata and offer discovery services, and secure cloud-based “Sandboxes” support multi-party data analysis. This proven infrastructure design provides a blueprint for other projects that need to analyse large amounts of heterogeneous data.

## BACKGROUND

Solve-RD is a Horizon 2020 supported EU flagship project that brings together >300 clinicians, scientists, and patient representatives from 51 institutes across 15 countries [1]. Solve-RD is built upon a core group of four European Reference Networks (ERNs) ERN-ITHACA, ERN-RND, ERN-Euro NMD and ERN-GENTURIS) and two associated ERNs (ERN RITA and ERN-EpiCARE) that annually see more than 270,000 rare disease (RD) patients with varying pathologies. The main ambition of Solve-RD is to solve unsolved RD cases for which a molecular cause is not yet known. This is achieved through an innovative clinical research environment that introduces novel ways to organise expertise and data. Two major approaches are being pursued: (i) massive data reanalysis of >19,000 experiments (various forms of genetic testing) from individuals affected by a rare condition and their unaffected family members and (ii) combined analysis of diverse types of newly-generated data, (’novel’ omics data).

For the data reanalysis, ERN partners contributed pseudonymised data (phenotypic data, pedigree information, exome sequencing (ES) data / genome sequencing (GS) sequencing data and associated metadata) for individuals affected by a RD who remained genetically undiagnosed after ES or GS. Data were submitted via the RD-Connect Genome-Phenome Analysis Platform (GPAP) [2]. In addition, novel omics data (short- and long-read GS, short and long-read RNA-sequencing, epigenomics, metabolomics, and Deep-ES) are being generated by different service providers within cohorts defined by the Data Interpretation Task Forces (DATF) from the four core collaborating ERNs [1]. Sample submitters from the ERNs upload their pseudonymised phenotypic and pedigree information in the RD-Connect GPAP PhenoStore module. From there, Phenopackets and pedigree descriptions in PLINK PED format are exported and submitted to the European Genome-phenome Archive (EGA). When novel omics data is generated, the service providers upload it directly to the EGA together with a manifest that links it to the corresponding individual. With such an amount of data to be analysed in a collaborative manner, downloading and analysing on local compute facilities is not feasible for all centers. Therefore, also centralised analysis facilities were desired.

All this clearly highlights the project’s need for a supporting data infrastructure. In particular because diverse demographic, phenotypic and multi-omics data needs to be securely submitted by a large number of clinical centres and other data providers, over a multi-year period. The quality of data and the relationships between data and files need to be captured to enable optimal use of the available data. Furthermore, to enable researchers from different centers to work together on the same dataset a cloud infrastructure is needed accessible by all researchers.

To enable reproducibility of analyses we organised the datasets in freezes of fixed sets of participants, that were updated with patches containing new information that became available over time. This information is captured within a MOLGENIS database [3; 4] and supplemented with an advanced discovery layer based on Café Variome [5] to enable identification of cases or sets of cases (virtual cohorts) based on a wide array of filters, including phenotypic or genotypic similarity metrics and federation with other RD data and sample resources. Moreover, appropriate metadata (e.g. file checksum) is collected to ensure that file integrity is maintained during transfer between research centres. This allows researchers to select samples of interest, for instance all affected individuals with a specific phenotype, and collect the associated files at their preferred analysis location. Similar discoverability features are available through the RD-Connect GPAP cohorts application. Furthermore, the RD-Connect GPAP is connected to MatchMaker Exchange [Boycott et al, 2022] and the Network of Beacons [6], enabling bidirectional patient matchmaking queries to similar resources around the world.

This infrastructure has been constructed by leveraging existing data platforms, tools and standards wherever possible, and by creating new tailored implementations where necessary, assembled into a unified infrastructure. We have operated on the core principle that we will reuse, enhance and deploy existing solutions (for core analytics support, databasing, data discovery and data sharing) wherever possible, according to FAIR data principles [7]. This paper describes the current state of the infrastructure which is fully operational, and indicates how we are further improving and extending its capabilities to ensure its future relevance and wider utility. We believe the resulting infrastructure could provide a template which future large scale RD analysis projects can start from.

## RESULTS

The data infrastructure we developed for Solve-RD facilitates submission of input data, a common approach to processing and archiving, collaborative data analysis, and sophisticated data discovery. The overall design and data flow is summarised in Figure 1.

**Fig. 1:**
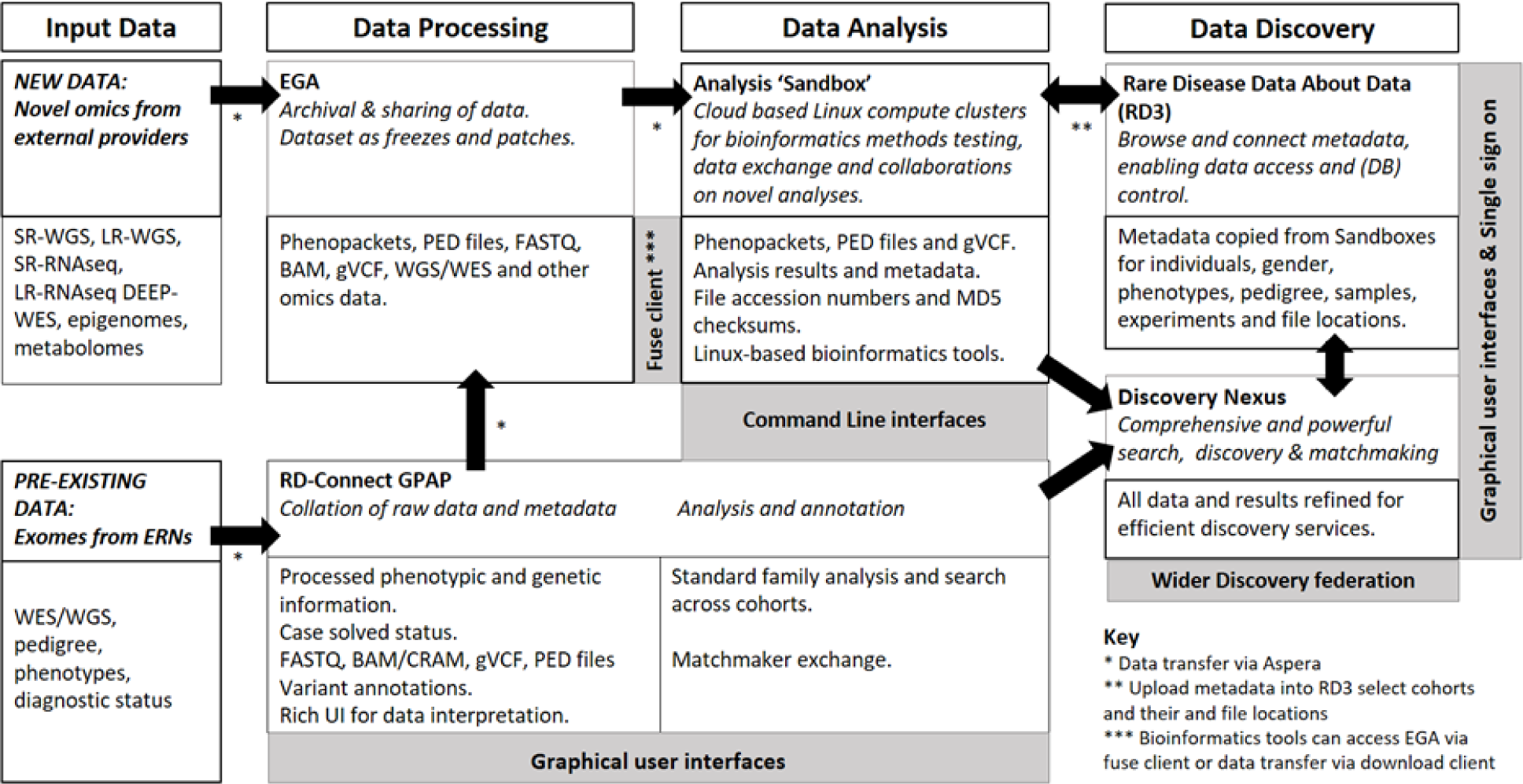
Rare disease analysis infrastructure overview. GS, genome sequencing. ES, exome sequencing. LR-GS, long-read genome sequencing. SR-GS, short-read genome sequencing. LR-RNAseq, long-read RNA-sequencing. SR-RNAseq, short-read RNA-sequencing. DEEP-ES, Deep sequencing ES. EGA, European Genome-phenome Archive. ERN, European Reference Network. GPAP, Genome-phenome analysis platform. UI, user interface. Although not depicted in the figure, the Solve-RD dataset is also discoverable through the participation of the RD-Connect GPAP in Matchmaker exchange and the Beacon Network.

### DATA SUBMISSION AND PROCESSING

Experimental metadata are first submitted to the RD-Connect GPAP, and corresponding phenotypic data submitted to the GPAP PhenoStore, where patient, phenotypic and family information are stored. Associated omics and pedigree data files then follow one of two paths, as described in the methods. Pre-existing sequencing data are submitted to the RD-Connect GPAP as FASTQ [8], BAM [9], or CRAM [10] files via a RedIris Aspera server. After processing, raw data, alignments whereupon they are processed and forwarded as BAM/CRAM and gVCF files to the EGA to be archived. Newly-generated novel omics data are directly archived to the EGA via a RedIris Aspera server. They are then downloaded by project partners and processed with a standard alignment and variant calling pipeline [11] to homogenise results and facilitate systematic analysis, interpretation, and comparisons. These processed data are submitted to the GPAP analysis platform.

At the EGA a unique file identifier is added to each individual file and data are made available for download. In parallel the Solve-RD Rare Disease Data about Data (RD3) database collects data and metadata on subjects, samples, experiments and files from these sources, and makes this available for discovery using the Discovery Nexus service, both described below.

### Standard Processing of reanalysis samples

Sequencing data originating from 43 different research centres was submitted together with a common set of required metadata for each participant and associated experiment. Solve-RD includes fully reanalysed ES or GS data from 22,326 participants (data freezes 1-3) for whom routine diagnostic procedures failed to achieve a molecular diagnosis. Furthermore, novel omics data from 5,184 participants (2,280 SR-GS, 510 LR-GS, 634 SR-RNAseq, 80 LR-RNAseq, 480 Epigenomics, 930 DEEP-ES, 270 Metabolomics) has been newly generated and incorporated. All of these data will be fully processed within the project [1; 12]. Solve-RD has archived over 750,000 files of primary and processed data at EGA totalling 818 terabytes. Impressively, this represents nearly 5% of all data archived at EGA, the second largest project at EGA to-date. The data held by the EGA will be fully available, under controlled access, to the wider RD community, and the ES/GS variant data is available to browse and analyse by any registered RD-Connect GPAP user.

### Long-term storage and file integrity

To ensure data security, data files are submitted to EGA in an encrypted format. accompanied by a manifest file (supplementary table 1). To ensure data integrity is preserved during file transfer and archival at EGA, file checksums are compared at different points of the submission process. For example, encrypted file checksums are compared before and after upload via Aspera to the EGA to ensure that the file was not corrupted during transfer. After being re-encrypted at EGA with a symmetric key and stored in the permanent archive, one final checksum check is performed to ensure integrity of the permanently archived, encrypted file.

### Interoperability

As described in our Methods section, the standard file formats used within our workflow, led to easy hand-off capabilities between the different components. Phenotypic and family information were stored in RD-connect GPAP using, respectively, the Phenopacket format [13] and the PLINK PED format [14; 15]. Next to the genomic data (FASTQ [8], BAM/CRAM [9; 10], and GVCF [16]) These were exported to the EGA and thereby given unique file identifiers, before being copied into the RD3 database to be accessible via the project Sandboxes.

### Freezes and patches

Data are structured into freezes and patches [1]. The Solve-RD project has generated three large freezes that consist of reanalysis data from subjects and experiments that have been submitted prior to one of three deadlines, meaning that each freeze consists of a fixed number of experiments and participants. The submission closing date for the first freeze was 30 September 2019 and it included data from 8,275 participants. The second and third freeze closed on 30 September 2020, and included data from 3,192 participants. The third freeze closed on 30 September 2021 and included data from 10,516 participants. A small fourth freeze is now being created, and currently contains data from 237 participants. Changes in data or metadata for these subjects are captured in patches, leaving the original dataset on which analyses have been performed intact, making reanalysis possible. In addition two data freezes for the novel omics are generated. For a small number of participants there were unintended duplications of datasets; a few cases had to be withdrawn from the collection for different reasons. To allow for data changes post-submission (e.g. addition of new phenotypic information or correction of errors), serial patches were introduced for each freeze. Patched files were released with a date inserted between the preserved filename and its file type extension (i.e., FILENAME.YYYY-MM-DD.extension). For each original freeze or subsequent patch all data was included in a uniquely identifiable EGA dataset (EGAD).

### DATA ANALYSIS

Data analysis was performed by data analysis task force (DATF) teams and interpretation of variants was done by data interpretation task force (DITF) teams. DATF activities were divided over several working groups [1] tackling ES and GS reanalysis and processing the newly generated ‘novel omics’ data. Only approved researchers who had signed the project code of conduct (Supplementary information 1) could access the data. Solve-RD partners are able to analyse data through three main approaches: the RD-Connect GPAP, a cloud-based ‘Sandbox’ and authorised local clusters.

While a wide range of analyses can be performed using the RD-Connect GPAP user interfaces (as described in the methods section) new analysis methods to find or interpret new variants and solve cases are continuously being developed. Moreover, for the novel omics data, analysis protocols are not yet standardised and needed to be developed by Solve-RD partners. We therefore needed an extensive analysis infrastructure to enable project analyses. A data request and download option was provided for partners that had their own substantial local compute facilities after approval of the project steering committee.

To support groups that did not have large compute and storage capacity, and also to enable multi-centre collaborative analyses, a centralised analysis ‘cloud’ Sandbox was established. The Sandbox is a high performance trusted research environment based on Linux. It supports existing and new research methods and also allows to collect and share project results. The Sandbox approach provides a central analysis environment for bioinformaticians to collaborate and to use and develop new methods freely. It applies strict access control procedures to ensure good governance and respect the trust given by data partners and the RD families whose data are being reanalysed. Via the Sandbox, DATF and DITF working groups can undertake pilot studies using newly devised tools to assess their added value, before undertaking an analysis of full datasets.

### Data management within analysis Sandbox

In addition to compute power and bioinformatics tools, we provide the Solve-RD Sandbox. The Sandbox functions as “Virtual/Trusted Research Environment” (VRE/TRE) or ‘Safe Haven’, providing access to data for analysis while protecting patient confidentiality supported by trained staff and agreed processes, see Kavianpour et al. [17] for a review. Before users could access any Sandbox content, a Consortium Code Of Conduct had to be signed and approved.

The Solve-RD Sandbox provides a Linux-based high performance compute (HPC) environment suited to bioinformaticians. To provide failover, we have deployed the Sandbox on two separate clouds, i.e., at Embassy (European Bioinformatics Institute) and at University of Groningen. The Sandbox supports large-scale data storage organised as a high-performance temporary (tmp) section and a stable but slower back-upped permanent (prm) folder. The tmp folder supports data analysis and so has a free structure for individual users to manage. The prm folder has a fixed structure that was identical at both Solve-RD Sandboxes.

Within each of the two VREs the tmp folder includes a single master folder containing original freeze files as well as patched files. For each freeze and patch a folder exists that carries symlinks to the files included in the specific patch release, typically a mix with the majority of files included in the previous patch and some new changed files. Because of limited storage space not all files from the project could be simultaneously held in the Sandbox. Therefore, larger files (BAM/CRAM) were omitted and reloaded as and when needed. In addition to these folders an ega-fuse-client folder was present in the prm folder, giving direct access to the Solve-RD datasets archived at the EGA. This enables BAM files to be accessed from within the VRE even though no local copy was present.

To provide access to analysis results, a dedicated directory was created for each DATF working group. To store their analysis results, each DATF working group appointed a data manager who was allowed to copy, move and remove data to and from the prm folders on the VREs (automatically synchronised between the two VRE instances). The folders were structured such that data sharing was optimally facilitated (Figure 2).

**Fig. 2:**
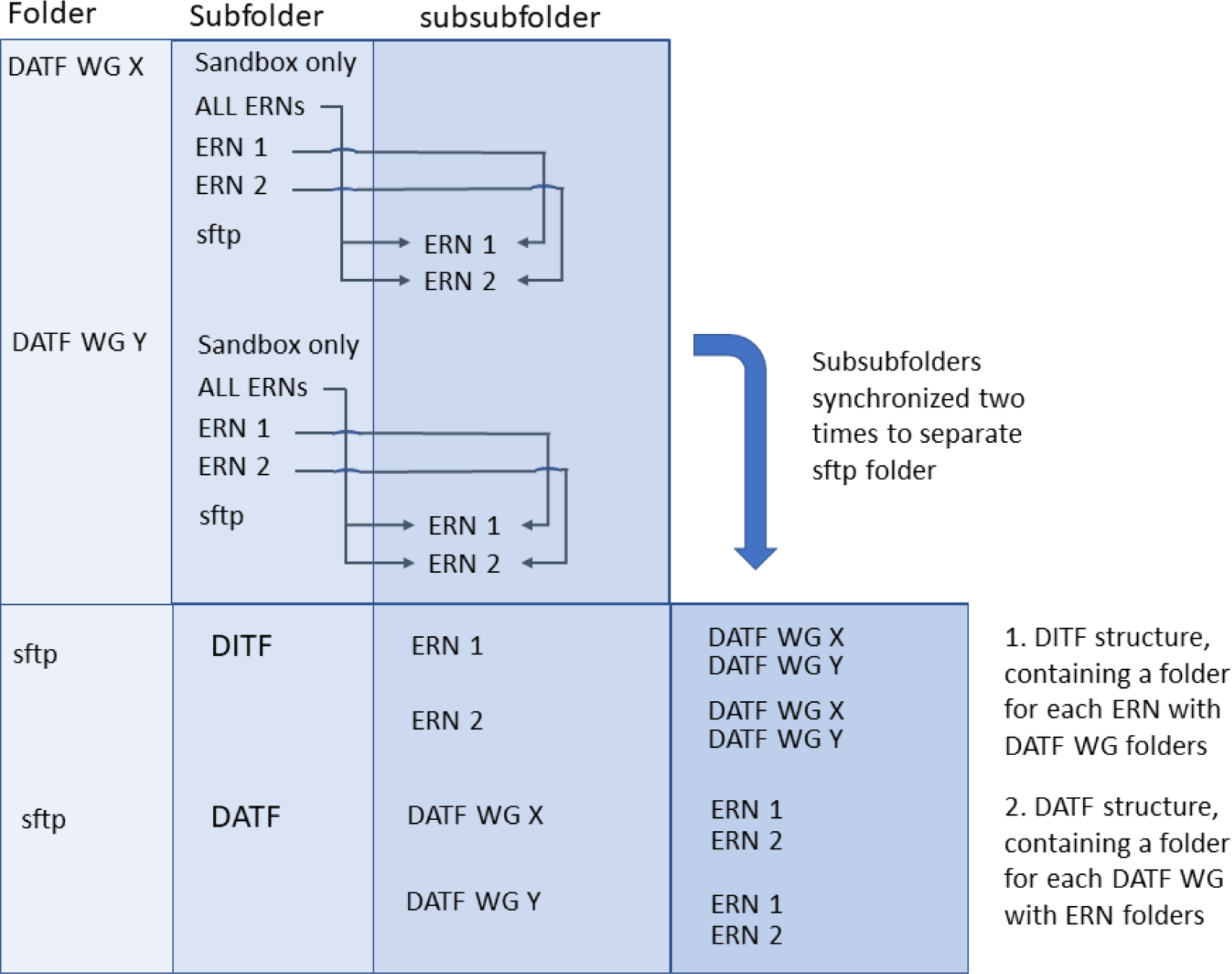
Sandbox folder structure. Data is organised by the data analysis working groups (DATF WG) in either folders per European Reference Network (ERN) or a common folder (for data intended for all ERNs). Additionally, large files that should be kept but not shared are stored in a ‘Sandbox only’ folder. All data to be shared with the ERNs is linked to an sftp folder with a subfolder per ERN accessible via SFTP access protocol. Thin arrows indicate links between specific subfolders. These folders are further synchronised to two folders: DATF and DITF (data interpretation task force), each with the same information (indicated by the thick arrow). The DATF folder has the same structure as the initial sftp folder (a folder for each DATF WG with subfolders per ERN). The DITF folder has the converse structure (a folder for each DITF ERN with subfolders per WG). This structure makes it easy for both DATF and DITF to browse the data (e.g. all CNV data or all data from ERN-ITHACA).

### DATA DISCOVERY

A large number of diverse data types and files exist within the Solve-RD project (multi-omics, variant interpretation, phenotyping, demographics, etc), and these are stored in different places and in different formats. The totality of metadata can be navigated via RD3 database, based on the MOLGENIS technology [3;4]. Based on RD3 bioinformaticians in the DATF can create inputs for their analyses. This works perfectly if one knows in advance what patients/samples/files one wants to select. However, there is often a need to find and select files based on the data values within them (e.g. based on specific variants in a VCF file [18]). Therefore, an advanced data discovery layer, called ‘Discovery Nexus’, was created on top of RD3, based on the Cafe Variome technology [5].

Additional data discovery functionalities are provided by the RD-Connect GPAP, as described in [2]. These consist of an internal “search across all” functionality, allowing users to search for specific types of variant in candidate genes of interest across all experiments. This can be further refined using the “cohorts” application which allows identification of affected individuals with similar phenotypes within the RD-Connect GPAP, including data not submitted as part of SolveRD. The RD-Connect GPAP is also an active node in the international MatchMaker Exchange network, facilitating patient matchmaking worldwide [19], and has also lit a beacon within the GA4GH Beacon Network [20].

### RD3 - tracking files and metadata

Direct data navigation is supported by the ‘rare disease data about data’ (RD3) system. This MOLGENIS database provides a complete listing of all patients/participants, samples, experiments and data files in Solve-RD, including EGA unique file identifiers. The many data types and relationships in Solve-RD are summarised in Figure 3.

**Fig 3:**
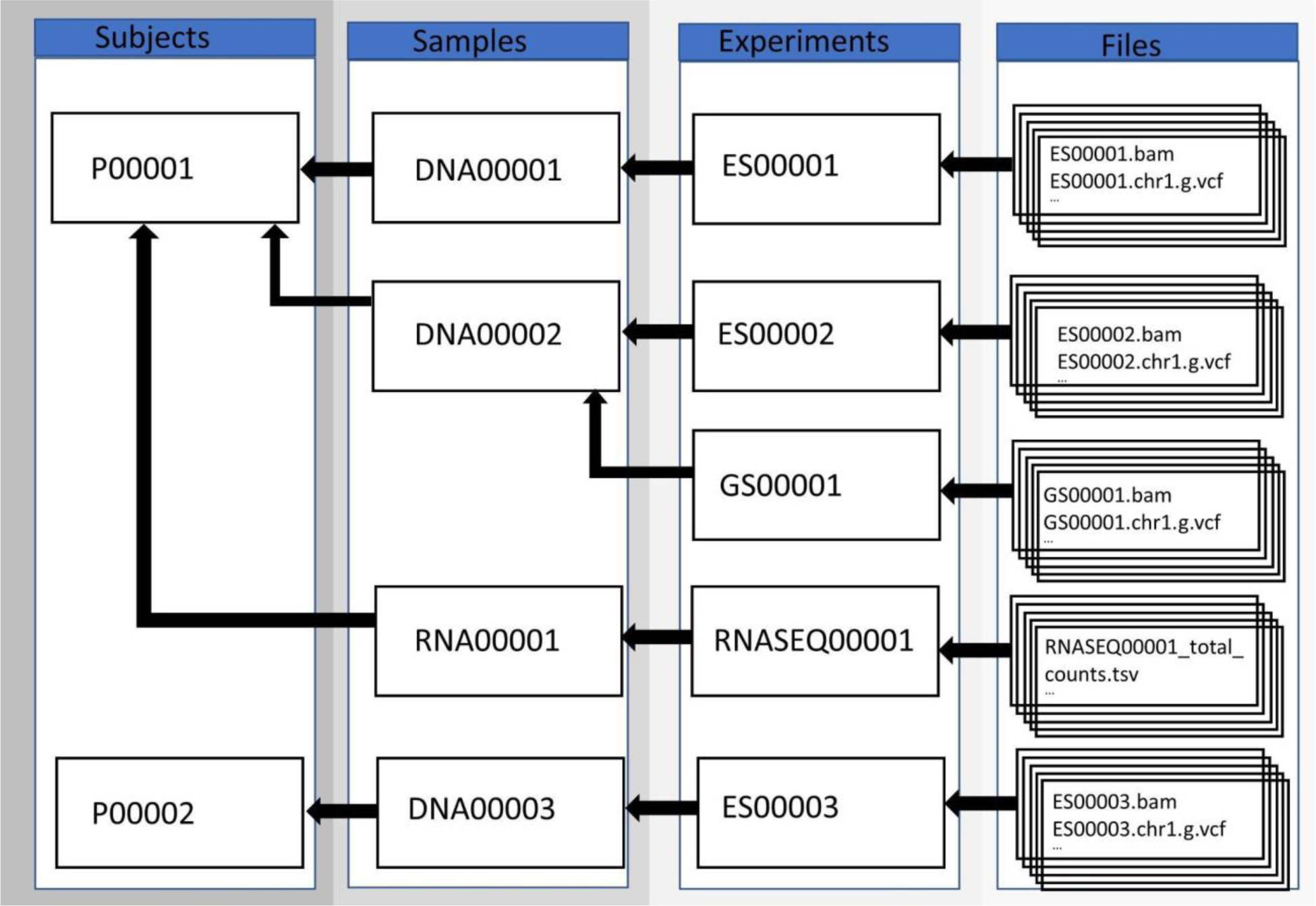
Data and metadata relations within Solve-RD. Arrows indicate the ‘derived from’ direction, e.g. Sample DNA00001 is derived from Subject P00001. We distinguish four main data/metadata types: subject, sample, experiments and files, with each derived from the former. This figure is actually a simplification as data is further organised in data releases we call ‘freezes’ and can be used in different combinations as ‘analyses’.

Some relationships are direct, such as the subject-sample relation (a sample is derived from a subject), whereas others are not so obvious and need to be discovered. RD3 is tightly integrated with Discovery Nexus, which also leverages useful extractions of various data files (e.g. extant variants, their frequency, host gene, mutation type, etc). Following a successful Discovery Nexus search, suitably permissioned users can click through to RD3 directly to access the discovered data files.

### Discovery Nexus

Discovery Nexus supports data discovery via a range of approaches that help users initially establish the existence and location (rather than the substance) of data within the system. The interface provides filtering options by which users can distil a comprehensive overview of selected datasets that might be of value for their intended purpose. Querying by multiple data values is possible, driven by ontologies and ontology cross-mappings. Searches can look for identity or semantic similarity to an entered term, or any combination of terms, and even extends to bridging between concepts (e.g., searches by biochemical pathway leverage knowledge of which genes are in each pathway). It also supports the GA4GH standard Beacon-2 API [6] for wider interoperability.

## Discussion

To enable a large number of researchers and clinicians to work together on a large dataset in Solve-RD, it was essential to establish a good data infrastructure. The solution we created includes access policies and procedures, including the code of conduct (supplementary information 1), a network of databases, HPC clusters, long-term storage capabilities, federated discovery services, tools and pipelines to provide the project with the ability to solve many RD cases that had not been solved using conventional strategies. Using this infrastructure, the Solve-RD project has already made >500 new diagnoses were achieved within the Solve-RD project [12], and many analyses powered by novel omics data are still ongoing.

The two parallel tracks, reanalysis of existing ES or GS data and novel omics data analysis, each created distinct challenges. One of the main challenges of the exome reanalysis stemmed from the heterogeneity of the submitted data. Cases were provided by institutions all around Europe and exomes were enriched using various designs and versions, and sequenced using different short-read platforms, each of which will result in different biases. In addition, analyses prior to submission to Solve-RD had been performed using a range of different alignment and variant-calling algorithms. For this reason, the Solve-RD project reanalysed primary sequence data from the earliest possible point, thereby eliminating bioinformatic-related differences and providing a coherent set of files for each of the experiments submitted. In parallel, the RD-Connect GPAP processed participant metadata and pedigree information and exported these in standard file formats. This provided reusable and interoperable data enabling downstream analysis via the RD-Connect GPAP, the project Sandboxes and local clusters.

Regarding novel-omics the main challenges from the perspective of the infrastructure were the different types of files produced and differences in accompanying metadata, which required a custom-made database format to capture this data.

Data FAIRness was enhanced by placing the data within the EGA data archive for long-term storage, request and access. To maximise user convenience, single-sign-on capability was provided across different components supporting a single goal, such as RD3 and Discovery Nexus, or between the Sandboxes and EGA via the filesystem in userspace (FUSE) client, as described in the methods. We also developed innovative methods to make data findable before and after data access is granted, using Discovery Nexus for preliminary searches (interoperable with GA4GH Beacon technology), and the RD3 database for full dataset navigation. Once the Solve-RD funding period is over, this same service will enable ERN data owners to advertise their data to researchers outside the project without directly releasing data too liberally or before access requests are reviewed and data sharing agreements set up. The data discovery service will also provide potential users with sufficient insight into the nature of available datasets to be confident that it is worth investing effort to request and analyse the data.

Within projects such as Solve-RD, concrete analyses are often conceived after the collection of data. This reflects the continuous expansion of associated knowledge and support tools. To facilitate this, we emphasised structured collection of rich metadata, thereby making the available data unambiguous in terms of its scope, quality, provenance and location. RD3 was used to organise and provision these metadata, following FAIRGenomes guidelines [21]. In addition, the RD-Connect GPAP co-hosts sections of the metadata relevant to their content, and this metadata also allows cohort-building via both Discovery Nexus and the RD-Connect GPAP.

In conclusion, Solve-RD has devised, implemented and validated an infrastructure for bringing together a set of reusable tools and best practices. As Solve-RD partners continue to use the infrastructure to perform many multi-omics analyses, the operational support teams are actively synergising with related projects. For example, some of the component are being deployed and expanded in European projects such as the European Joint Programme on Rare Diseases (EJP-RD, https://www.ejprarediseases.org), the EU Genome Data Infrastructure project (GDI, https://gdi.onemilliongenomes.eu/), and national initiatives such as the Dutch FAIR genomes/Health-RI genomics project (https://www.health-ri.nl/). Hence, the infrastructure described in this paper can be used as a blueprint for future multi-omics data (re)analysis projects and data hubs.

## Methods

### DATA SUBMISSION AND PROCESSING

Many types of data were provided by the ERNs or newly generated within the Solve-RD project, including demographic and phenotypic data of participants and metadata on samples, experiments and files. Pre-existing sequencing data are submitted to the RD-Connect GPAP as FASTQ [8], BAM [9], or CRAM [10] files via a RedIris Aspera server. Specifically, ES and GS reanalysis data and metadata was provided by partners of six different ERNs: ERN-ITHACA, ERN-RND, ERN-Euro NMD, ERN-GENTURIS, ERN RITA, ERN-EpiCARE. For novel omics analysis, various other file types and concomitant metadata were produced.

Raw ES and GS read data for reanalysis, together with accompanying metadata and deep phenotypic descriptions of affected individuals were submitted by Solve-RD partners to the RD-Connect GPAP (GPAP). Alignment and short variant calling was undertaken for all experiments using an an identical variant calling workflow, in order to minimise bioinformatics induced artefacts, as described in [11], all identified SNVs and InDels were made immediately available to Solve-RD collaborators for analysis in the GPAP Genomics module. Subsequently the raw data, and processed data in the form of BAM/CRAM and gVCF files were transferred to the European Genome-phenome Archive (EGA) for longer-term archival and redistribution to other Solve-RD partners.

The RD-connect Genome-Phenome Analysis Platform (GPAP) was used for collation of all phenotypic data, and standardised processing of all short-read ES and GS data submitted to Solve-RD. Data collation was undertaken as described in Laurie et al., 2022 [2].

Briefly, in the first step pseudonymised phenotypic descriptions of all affected individuals were uploaded to the RD-Connect GPAP PhenoStore module, using HPO, OMIM and Orphanet terms to generate a detailed phenotypic description, together with a family tree linking individuals. Each individual receives a unique participant ID (P-ID) and for accompanying experiments E-IDs (experiment IDs) were created. In the second step, metadata describing the raw sequencing data to be submitted for reanalysis, and linking it to the individual’s phenotypic record is uploaded to the GPAP Data Management module. Finally, the raw sequencing data itself is transferred using a robust, high-speed Aspera data transfer service provided by RedIris (https://www.rediris.es/), the Spanish academic and research network. Once submission is complete, the data is automatically imbibed and processed by the automated standard analysis pipeline.

### Standard analysis pipeline

For joint data analysis, it is important that technical differences between experiments are minimised. Therefore, using the CNAG-CRG local HPC resources, all short-read ES and GS data submitted to Solve-RD were reprocessed using an identical standardised variant calling pipeline as described in Laurie et al., 2016 [11].

### Data sources for pre-existing and new data

For reanalysis of ES/GS, novel omics Short read (SR)-GS, and DEEP-ES data, the starting point for reanalysis was the associated FASTQ files. When BAM or CRAM files were submitted, these were first transformed back to FASTQ. Using the standard analysis pipeline (Figure 1), data were processed in a standardised manner as described above, producing a single BAM and 25 g.VCF files (autosomes, X, Y and MT), accompanied by .BAI and .TBI index files, respectively. Phenotypic information was exported from GPAP in Phenopacket format and pedigrees in PED file format. LR-GS files and RNA-sequencing data were stored in BAM format. Data analysis produced output of various file formats, depending on the tools used for analysis.

### Interoperability

To maximise interoperability for tool integration and reuse beyond Solve-RD and to overcome language barriers, we use widely adopted and machine-readable international and community standards and ontologies whenever possible. Within PhenoStore, deep phenotypic descriptions are recorded using Human Phenotype Ontology [22], Orphanet Rare Disease Ontology (ORDO: https://www.orphadata.com/ontologies/) [23] and the Online Mendelian Inheritance in Man (OMIM) [24] terminology. Phenotypic records can be exported using the GA4GH approved Phenopacket format [13], and family trees in PLINK PED format [14; 15]. Genomic alignments are stored and transferred (e.g., to the EGA) in GA4GH approved BAM, CRAM formats (https://www.ga4gh.org/genomic-data-toolkit/). Variants are stored in GVCF format [16]. Biological annotations, available in the Data Analysis module are provided by Ensembl VEP [25], and supplemented with data from other genomics community resources such as ClinVar [26], gnomAD [27], and PanelApp [28]. Data discovery and sharing is achieved through the implementation of GA4GH Beacon-V2 [6], and MME APIs [19]. Partner involvements in other initiatives also guided our work regarding other standardisation strategies, not least B1MG, GA4GH, FAIR genomes [21], ELIXIR, BBMRI and EJP-RD.

### EGA long-term data archiving and access

The European Genome-phenome Archive (EGA) (https://ega-archive.org/) is a service for permanent archiving and sharing of identifiable genetic and phenotypic data [29; 30]. Data archived at the EGA ensures long term availability, interoperability, and identifiability during projects and beyond. The primary objects in the EGA data model are studies, datasets, and files (raw and processed). Each archived file is assigned an EGA accession functioning as a unique identifier (UID). Moreover, each file can be part of one or more datasets, each with its own accession number. After data are successfully archived and released, the EGA provides access to the data only upon approval by the associated Data Access Committee (DAC) for specified individuals. Datasets can be accessed using the PyEGA3 streaming client (https://github.com/EGA-archive/ega-download-client) and a filesystem in userspace (FUSE) client (https://github.com/EGA-archive/ega-fuse-client).

To ensure data are FAIR, metadata are uploaded to EGA alongside data files. These metadata take the form of manifest files (supplementary table 1) which contain many attributes describing the data, for example what library preparation and sequencing strategy was followed, what type of data analysis was done including which reference genome was used, and minimal public information about the study subjects. Manifest files are converted to the EGA XML standard for representing metadata before being permanently archived.

## DATA ANALYSIS

### RD-Connect GPAP

The RD-Connect GPAP allows users to perform variant analysis to identify potential disease-causing variants in a single proband or any family structure, and allows user-defined queries across a cohort of affected individuals. These capabilities are provided via a user-friendly interface suitable for clinicians, genome scientists and bioinformatics researchers.

A large variety of filters can be applied in order to identify known pathogenic variants e.g. described in ClinVar, or prioritise variants that are potentially pathogenic for further investigation [2]. Furthermore, variants can be visualised in remotely hosted native BAM files on-the-fly, directly within the GPAP, through implementation of the GA4GH htsget streaming protocol and a client-side Integrative Genomics Viewer instance [31].

Analysis can be undertaken in two different ways, either interactively via a graphical user interface (GUI) or automated via a Python-based API. The interactive approach is ideal for analysing individual families and applying different filter strategies. For processing large numbers of experiments, as undertaken in Solve-RD, programmatic batch analysis can be undertaken as described previously [32].

Intra-GPAP case-matching is possible via an instance of the GA4GH MME API (https://github.com/ga4gh/mme-apis) and by searching across cohort functionalities. External case matching can be achieved through the global MME API [19], and single variants can be found via the Beacon-V1 API [20].

### Sandboxes for bespoke bioinformatics analyses

Bioinformatics methods often require a Linux command-line environment and extensive computing and storage capabilities. In line with this, we implemented two Sandboxes as Linux-based HPC clusters that can be remotely accessed and act as a VRE/TRE. To enable reproducibility and reusability (i.e. in future projects) these Sandboxes are implemented as a ‘cloud’ service that can be automatically deployed at different cloud providers using the same playbook (https://github.com/rug-cit-hpc/league-of-robots), using OpenStack for virtualisation of Linux CentOS7 (https://www.centos.org) with Spacewalk (https://spacewalkproject.github.io) for package distribution and management and using the LMOD module system (https://github.com/TACC/Lmod) and Easybuild (https://github.com/easybuilders/easybuild) to reproducibly install bioinformatics tools.

Because HPC systems typically need large maintenance windows where the service is offline, we have two separate Sandbox installations at different locations to prevent a single point of failure and ensure continuous operations to the partners: one at EMBASSY (http://www.embassycloud.org/) [32], hosted by the EMBL European Bioinformatics Institute (EMBL-EBI), which has close connections to the EGA, and one at the University of Groningen Centre for Information Technology (https://www.rug.nl/society-business/centre-for-information-technology/) [33] attached to the University Medical Centre Groningen. The EMBASSY VRE is only accessible by members of the Solve-RD project, while the UMCG VRE is a larger facility shared with other projects beyond Solve-RD. A dedicated Solve-RD group is present in the UMCG VRE with access restricted to Solve-RD members only. The EMBASSY VRE has 40 Tb of storage and 12 compute nodes with 14 cores/node and 56072 Mb RAM/node. The UMCG VRE (http://docs.gcc.rug.nl/gearshift/) has shared storage with other projects, with 200 Tb reserved for the Solve-RD project and a total of 10 compute nodes available with 22 cores/node and 205490 Mb RAM/node.

### Access to analysis results

Both clusters use internal networks that are not directly accessible from the internet. Access is possible via dedicated jumphosts, security hardened machines not involved in any data storage or processing. Using asymmetric cryptography via a private-public key pair [34], users can login to the jumphost to be directly redirected to the main HPC cluster. To allow for data access for non-bioinformaticians, we created an SFTP transfer server that could be accessed using a graphical user interface such as WinSCP (https://winscp.net), MobaXTerm (https://mobaxterm.mobatek.net) or Cyberduck (https://cyberduck.io) via a private-public keypair without the extra security of a jumphost.

## DATA DISCOVERY

### MOLGENIS RD3

To manage metadata on subjects, samples, experiments and files of ES reanalysis and novel omics, we used the MOLGENIS RD3 database. A specific Solve-RD instance of this was created (https://github.com/molgenis/RD3_database), accessible via a web-interface (https://solve-rd.gcc.rug.nl/). In this database, metadata (e.g. file accession numbers) and data (e.g. average coverage for ES targets) are collected for all Solve-RD subjects and the associated samples, experiments and files.

Content includes information on how samples were collected and the subjects they came from, as well as the analyses that were performed and the location of the files generated. RD3 acts as a hub for GPAP data on Solve-RD participants, data provided by the EGA, files located in the Sandbox, and metadata required for the Discovery Nexus tool. Using portal tables, relevant data and metadata are imported into RD3 using a manifest file provided by the EGA.

RD3 was built in MOLGENIS [3,4], an open-source database platform for storing, managing, analysing, and sharing data. Approved users can log in using a local login or through FusionAuth (https://fusionauth.io/). All the relevant metadata for the research is collected within the Solve-RD RD3. The core structure of RD3 consists of several tables matching the different types of information that should be selected (Figure 3). ES reanalysis data was imported into RD3 using a system of freezes and patches. Each of these sections has the same format.

The subjects table contains information on the participants as collected in GPAP PhenoStore, imported via phenopackets and PED files archived at the EGA. Subjects are identified based on their P-ID. For each subject the P-IDs of the parents are given if they were included in the project, as is the family number to identify all subjects who are part of the same family. Furthermore, the subject’s sex and a disease name or the phenotypes known to be present (or absent) are listed. For each subject, it is recorded if they are considered to be affected by a condition or not (e.g. a child is affected and both parents are unaffected by a condition). In addition, information is stored on the case submitter, e.g. if they are allowed to be recontacted in case of incidental findings or if the case is retracted. Finally, the subjects table shows if the sample is solved. Because this information is updated in the GPAP PhenoStore, a connection between the two programs allows the solved status to be updated daily.

Zero or more samples may be derived from each subject. Sample metadata is collected in the samples table. Each sample is given a sample-ID (S-ID) for unique identification. Per S-ID, the P-ID of the subject from which it is derived is shown as well as the tissue type (e.g. whole blood) and other sample specifications.

Zero or more experiments can be performed on each sample (e.g. ES on DNA isolated from the sample). Information on these experiments is collected in the experiments table (see Figure 4). Each type of experiment has its own specific lay-out. For ES the enrichment kit used is captured as is the sample preparation method. The metrics “% of the target covered >20x“ and “average target coverage“ are also collected.

**Fig. 4:**
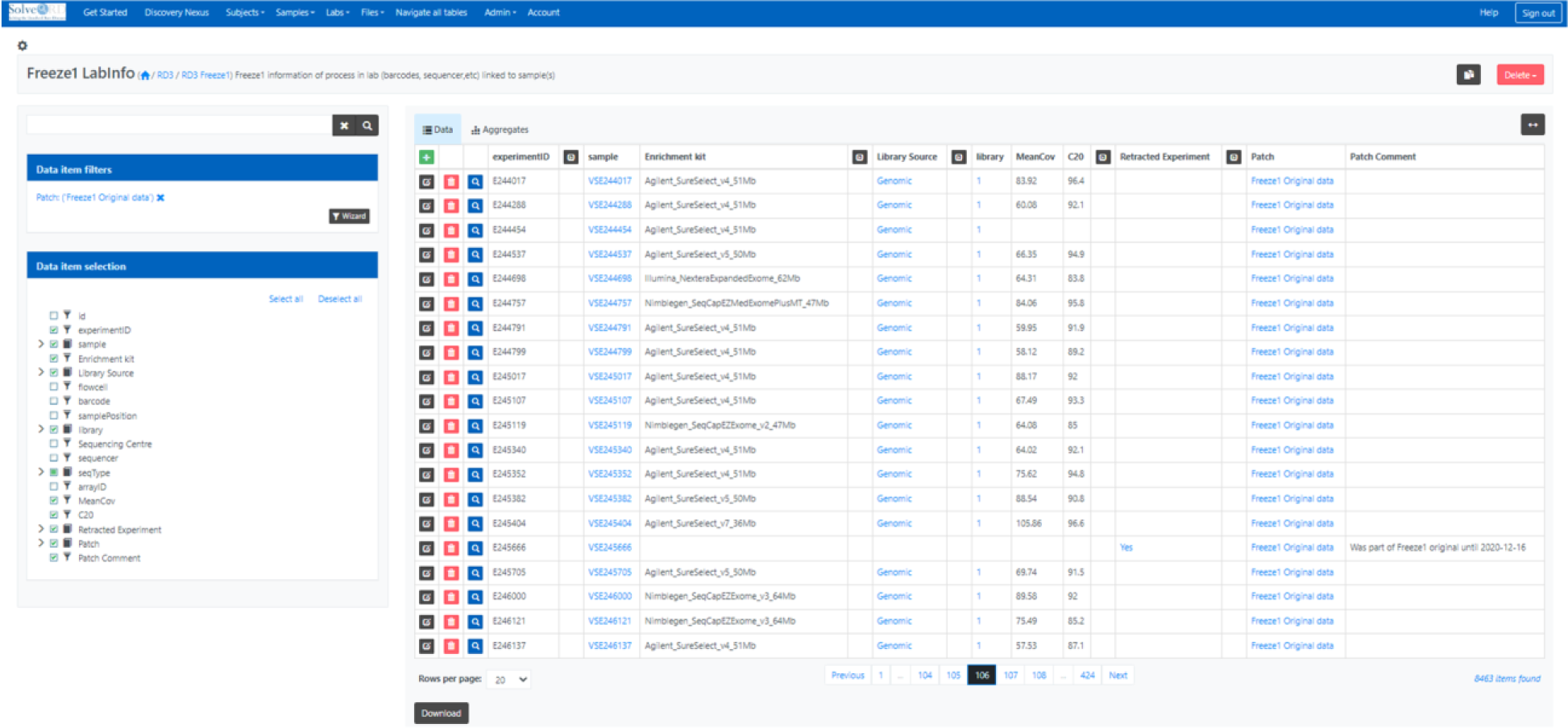
Solve-RD RD3 LabInfo screen showing a subset of the Freeze1 experiment data. On the left entries are filtered on patch ‘Original data’ and columns are filtered on interest. In the current view, the experimentID is connected to the sample on which the experiment was performed. In addition, information on the experiment is shown. For these samples genomic data was the input for exome sequencing experiments on which various different enrichment kits were used. For most of the samples statistics on the average target coverage (MeanCov) and number of bases covered by at least 20 sequencing reads (C20) was available. If a subject was retracted from the project, all metadata except identifiers were removed from the database and the experiment was labelled as retracted.

For each family, subject and experiment files are archived at the EGA. RD3 captures this information in the files table. Here, for each file, the path in the Sandbox and the VRE ega-fuse-client within the dataset are given with its checksum information enabling a sanity check on copies of this file. Information is recorded about the filetype, the experiment it belongs to and the EGA accession number.

### Discovery Nexus

RD3 is seamlessly integrated with Discovery Nexus using a single sign-on option based on the open ID connect protocol (OIDC, implemented using FusionAuth), which is compatible with the life sciences AAI, previously known as ELIXIR AAI [35], which we plan to implement in the future. The latter will enable users to sign in using their institute sign in, which increases security and GDPR compliance and ensures removal when contracts terminate.

Discovery Nexus is a parallel component to RD3 that provides advanced and more powerful capabilities for quickly and deeply searching Solve-RD data stored in different locations and formats. Built on Café Variome [5], Discovery Nexus abstracts direct database-style queries to concept-based queries, for example, phenotypes and diseases are based on common ontologies that Discovery Nexus dynamically maps to ontologies and hierarchies within ontologies used in the underlying subject phenotyping. This is also extended to querying using semantic similarity between and across ontologies. This abstraction allows Discovery Nexus to represent searches in an intuitive query builder interface focussed on elements that make queries based on demographics, phenotypes, diseases, variants, biochemical pathways, mutation characteristics, solved-or-not status, and data availability (Figure 5). This separation of query from database language also provides protection to subjects and studies identification as the actual data is not queried or represented in the query or results. For example, variants are not directly queried in Discovery Nexus; instead, the query interface allows searches for types of variant mutations in genes or gene families.

**Fig. 5A:**
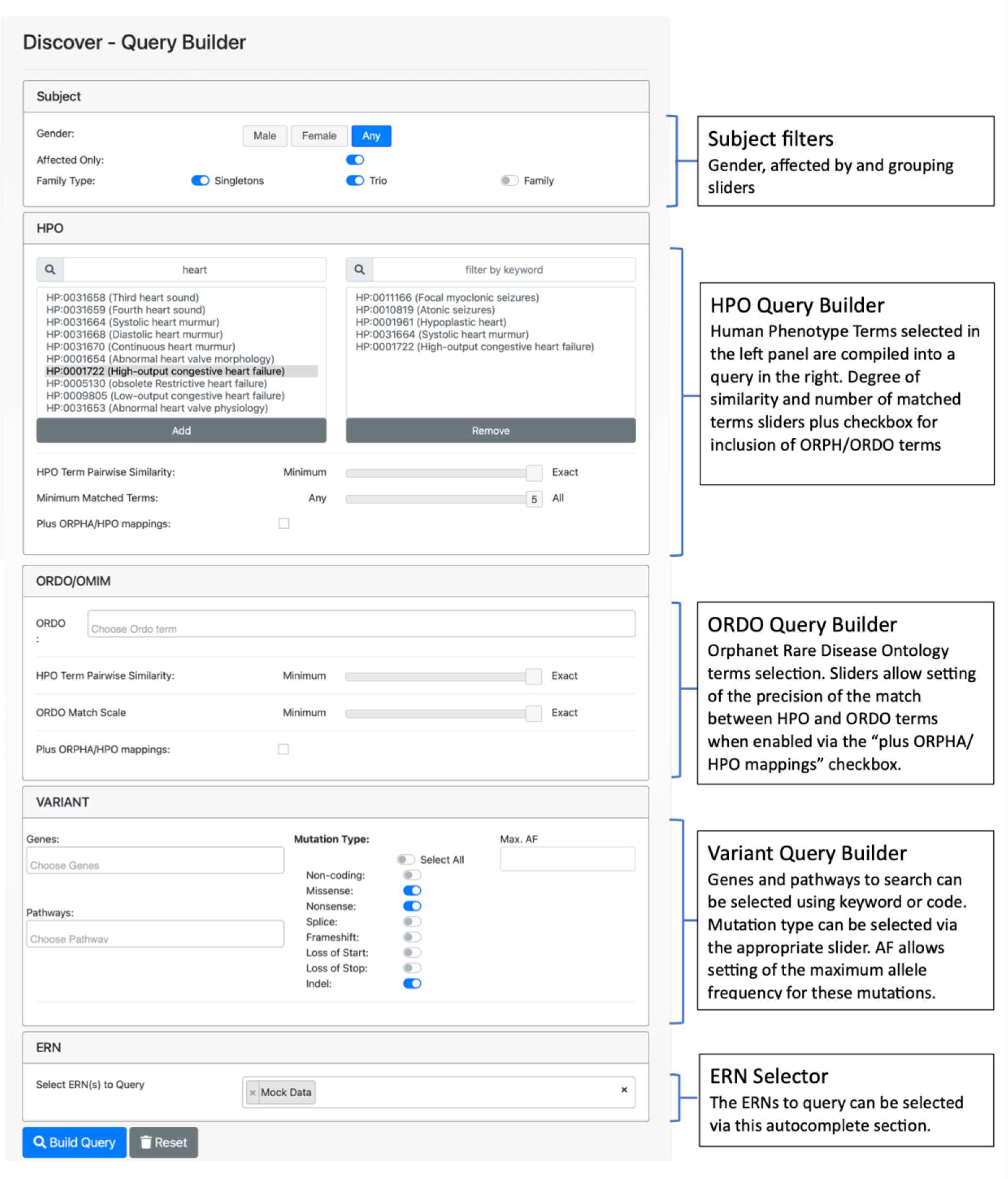
Discovery Nexus query interface. This interface supports querying by any combination of various demographic and inheritance (Subject Filters), phenotypes (HPO Query Builder), diseases (ORDO Query Builders) or suspected variant filters (Variant Filter). In the HPO Query Builder typing any part of an HPO phenotype term or code creates a visible list of relevant items to select from, whereupon they are transferred into the adjacent panel to form part of the query. Phenotype matching can specify matching on identical terms only (exact) or recover similar terms (based on a precomputed matrix of relationship scores and the position of the slider). The minimum number of matching terms can also be specified, creating an “OR” query, settings above the minimum creates a query that returns results that match at least the specified number of terms in any combination. HPO queries can also be instructed to interrogate phenotype data stored as ORDO terms. Matching of HPO to ORDO terms (in the ORDO Query Builder) is controlled by the use of the HPO pairwise similarity slider, to define the number of HPO terms that should match an ORDO term as well as the ORDO match scale, defining the specificity of the HPO term(s) to the selected ORDO term (based on a pre-computed matrix of their occurrence across all ORDO terms). Hence, when mapping ORDO to HPO terms, exact matching will traverse the mapping of these two term sets to find fewer but more specific HPO terms, while minimum matching will include more HPO terms but these may match other ORDO terms as well. Variant data cannot be filtered at the specific base-change level (as this would raise privacy concerns), but is instead queryable by host gene, allele frequency and mutation type using the Variant Query Builder. It is also possible to filter for variants based on affected biochemical pathways, given known relationships between genes and pathways (using the Reactome Knowledge base [36]). Finally, the ERN dataset to be queried must be explicitly stated and requires that the user has permission to query the specified ERNs.

**Fig. 5B:**
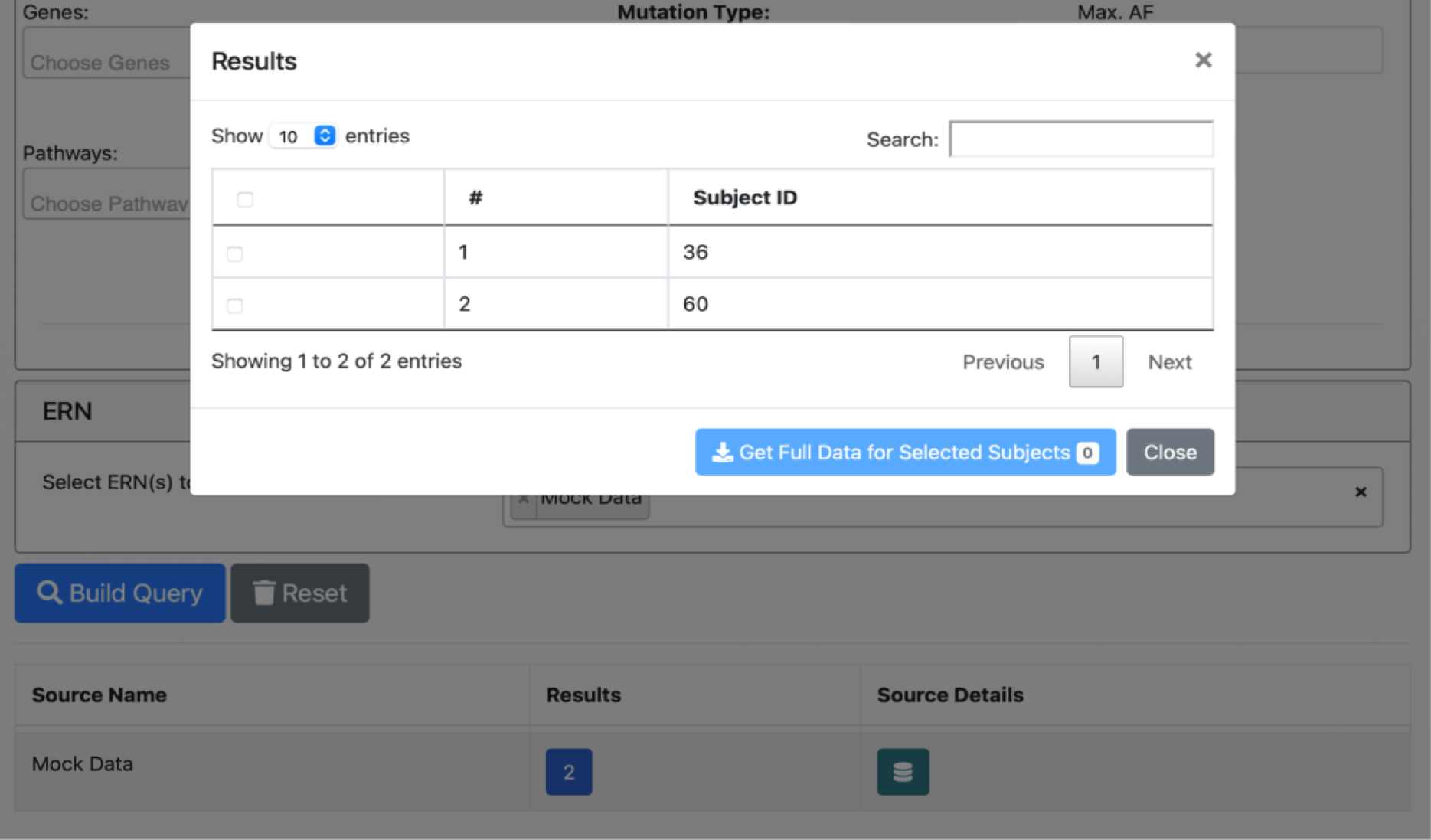
Discovery Nexus Query Results. After submitting the query using the “Build query button” the system will return a count for matching results in the resources selected. Clicking on the number in the blue box will bring up the summary pop-up window as shown above, giving basic details of the matches (again subject to the user having been assigned permissions). The blue “Get Full Data for Selected Subjects” will open a link to request access from the resources holding the required data (where this is available). Alternatively, clicking the green button in the source details, will open a summary page with contact details for the resource, where a direct link to request the data is not available.

### Handoff from Nexus to RD3 to get data

Discovery Nexus and RD3 operate under a federated single sign-on for authentication using the industry standard OIDC provided by RD3, with only users authorised by Solve-RD able to access either application. This allows the two parallel systems to interoperate seamlessly, with a handoff facility allowing search results in Discovery Nexus to be pre-populated in RD3, so that the user access information about the underlying data without logging in again.

### Data availability

Data will be deposited at EGA. Accession numbers to be provided. Pseudonymised phenotypic information for all individuals and their genetic variants are accessible through the RD-Connect GPAP (https://platform.rd-connect.eu/) upon validated registration. All raw and processed data files will be made available at the EGA (Solve-RD study EGAS00001003851) upon approval by data access committee. The Ethics committee of the Eberhard Karl University of Tubingen gave ethical approval for this work.

### Supplementary data

Supplementary table 1: EGA manifest file

Supplementary information 1: Solve-RD Code of Conduct.

## Supporting information

Supplementary information 1: Code of Conduct

Supplementary table 1: manifest file

## Acknowledgements

We acknowledge all Solve-RD partners (see Solve-RD consortium) and all hospitals and patients that shared data.

## Corporate author lists

### Solve-RD consortium

**EKUT:** Olaf Riess^1, 2^, Tobias B. Haack^1^, Holm Graessner^1, 2^, Birte Zurek^1, 2^, Kornelia Ellwanger^1, 2^, Stephan Ossowski^1, 3^, German Demidov^1^, Marc Sturm^1^, Julia M. Schulze-Hentrich^1^, Rebecca Schüle^1, 2^, Jishu Xu^4, 5^, Christoph Kessler^4, 5^, Melanie Kellner^4, 5^, Matthis Synofzik^4, 5^, Carlo Wilke^4, 5^, Andreas Traschütz^4, 5^, Ludger Schöls^4, 5^, Holger Hengel^4, 5^, Holger Lerche^1^, Josua Kegele^6^, Peter Heutink^4, 5^

**RUMC:** Han Brunner^7-9^, Hans Scheffer^7, 8^, Nicoline Hoogerbrugge^7, 10^, Alexander Hoischen^7, 10, 11^, Peter A.C. ‘t Hoen ^10, 12^, Lisenka E.L.M. Vissers^7, 8^, Christian Gilissen^7, 10^, Wouter Steyaert^7, 10^, Karolis Sablauskas^7^, Richarda M. de Voer^7, 10^, Erik-Jan Kamsteeg^7^, Bart van de Warrenburg^8, 13^, Nienke van Os^8, 13^, Iris te Paske^7, 10^, Erik Janssen^7, 10^, Elke de Boer^7, 8^,Marloes Steehouwer^7^, Burcu Yaldiz^7^, Tjitske Kleefstra^7, 8^

**University of Leicester**: Anthony J. Brookes^14^, Colin Veal^14^, Spencer Gibson^14^, Vatsalya Maddi^14^, Mehdi Mehtarizadeh^14^, Umar Riaz^14^, Greg Warren^14^, Farid Yavari Dizjikan^14^, Thomas Shorter^14^

**UNEW:** Ana Töpf^15^, Volker Straub^15^, Chiara Marini Bettolo^15^, Jordi Diaz Manera^15^, Sophie Hambleton^16^, Karin Engelhardt^16^

**MUH:** Jill Clayton-Smith^17, 18^, Siddharth Banka^17, 18^, Elizabeth Alexander^18^, Adam Jackson^17, 18^

**DIJON:** Laurence Faivre^19-23^, Christel Thauvin^19-23^, Antonio Vitobello^21^, Anne-Sophie Denommé-Pichon^21^, Yannis Duffourd^21, 22^, Ange-Line Bruel^21^, Christine Peyron^24, 25^, Aurore Pélissier^24, 25^

**CNAG-CRG:** Sergi Beltran^26, 27^, Ivo Glynne Gut^26, 27^, Steven Laurie^26^, Davide Piscia^26^, Leslie Matalonga^26^, Anastasios Papakonstantinou^26^, Gemma Bullich^26^, Alberto Corvo^26^, Marcos Fernandez-Callejo^26^, Carles Hernández^26^, Daniel Picó^26^, Ida Paramonov^26^, Hanns Lochmüller^26^

**EURORDIS:** Gulcin Gumus^28^, Virginie Bros-Facer^29^

**INSERM-Orphanet:** Ana Rath^30^, Marc Hanauer^30^, David Lagorce^30^,Oscar Hongnat^30^,Maroua Chahdil^30^,Emeline Lebreton^30^

**INSERM-ICM:** Giovanni Stevanin^31-35^, Alexandra Durr^31-34, 36^, Claire-Sophie Davoine^31-35^, Léna Guillot-Noel^31-35^, Anna Heinzmann ^31-34, 37^, Giulia Coarelli^31-34, 37^

**INSERM-CRM:** Gisèle Bonne^38^, Teresinha Evangelista^38^, Valérie Allamand^38^, Isabelle Nelson^38^, Rabah Ben Yaou^38-40^, Corinne Metay^38, 41^, Bruno Eymard^38, 39^, Enzo Cohen^38^, Antonio Atalaia^38^, Tanya Stojkovic^38, 39^

**Univerzita Karlova:** Milan Macek Jr.^42^, Marek Turnovec^42^, Dana Thomasová^42^, Radka Pourová Kremliková^42^, Vera Franková^42^, Markéta Havlovicová^42^, Petra Lišková^43, 44^, Pavla Doležalová^45^

**EMBL-EBI:** Helen Parkinson^46^, Thomas Keane^46^, Mallory Freeberg^46^, Coline Thomas^46^, Dylan Spalding^46^

**Jackson Laboratory**: Peter Robinson^47^, Daniel Danis^47^

**KCL**: Glenn Robert^48^, Alessia Costa^49^, Christine Patch^49, 50^

**UCL-IoN**: Mike Hanna^51^, Henry Houlden^52^, Mary Reilly^51^, Jana Vandrovcova^52^, Stephanie Efthymiou^52^, Heba Morsy^52^, Elisa Cali^52^, Francesca Magrinelli^53^, Sanjay M. Sisodiya^54^, Jonathan Rohrer^55^

**UCL-ICH**, Francesco Muntoni^56, 57^, Irina Zaharieva^56^, Anna Sarkozy^56^

**Universiteit Antwerpen**: Vincent Timmerman^58, 59^, Jonathan Baets^60, 61^, Geert de Vries^59, 60^, Jonathan De Winter^59-61^, Danique Beijer^58-60^, Peter de Jonghe^59, 61^, Liedewei Van de Vondel^58-60^, Willem De Ridder^59-61^, Sarah Weckhuysen^60, 62^

**Uni Naples/Telethon UDP**: Vincenzo Nigro^63, 64^, Margherita Mutarelli^64, 65^, Manuela Morleo^64^, Michele Pinelli^64^, Alessandra Varavallo^64^, Sandro Banfi^63, 64^, Annalaura Torella^63^, Francesco Musacchia^63, 64^, Giulio Piluso^63^

**UNIFE**: Alessandra Ferlini^66^, Rita Selvatici^66^, Francesca Gualandi^66^, Stefania Bigoni^66^, Rachele Rossi^66^, Marcella Neri^66^

**UKB**: Stefan Aretz^67, 68^, Isabel Spier^67, 68^, Anna Katharina Sommer^67^, Sophia Peters^67^

**IPATIMUP**: Carla Oliveira^69-71^, Jose Garcia-Pelaez^69, 70, 72^, Rita Barbosa**-**Matos^69, 70, 73^, Celina São José^69, 70, 72^, Marta Ferreira^69, 70, 74^, Irene Gullo^69-71, 75^, Susana Fernandes^76^, Luzia Garrido^75^, Pedro Ferreira^69, 70, 77^, Fátima Carneiro^69-71, 75^

**UMCG**: Morris A Swertz^78^, Lennart Johansson^78^, Joeri K van der Velde^78^, Gerben van der Vries^78^, Pieter B Neerincx^78^, David Ruvolo^78^, Kristin M Abbott^79^, Wilhemina S Kerstjens Frederikse^79, 80^, Eveline Zonneveld-Huijssoon^79, 81^, Dieuwke Roelofs-Prins^78^, Marielle van Gijn^79, 81^

**Charité**: Sebastian Köhler^82^

**SHU**: Alison Metcalfe^48, 83^

**APHP**: Alain Verloes^84, 85^, Séverine Drunat^84, 85^, Delphine Heron^86, 87^, Cyril Mignot^86, 88^, Boris Keren^86^, Jean-Madeleine de Sainte Agathe^86^

**CHU Bordeaux**: Caroline Rooryck^89^, Didier Lacombe^89^, Aurelien Trimouille^90^

**Spain UDP**: Manuel Posada De la Paz^91^, Eva Bermejo Sánchez^91^, Estrella López Martín^91^, Beatriz Martínez Delgado^91^, F. Javier Alonso García de la Rosa^91^

**Ospedale Pediatrico Bambino Gesù, Rome**: Andrea Ciolfi^92^, Bruno Dallapiccola^92^, Simone Pizzi^92^, Francesca Clementina Radio^92^, Marco Tartaglia^92^

**University of Siena**: Alessandra Renieri^93-95^, Simone Furini^93, 94^, Chiara Fallerini^93, 94^, Elisa Benetti^93, 94^

**Semmelweis University Budapest**: Peter Balicza^96^, Maria Judit Molnar^96^

**University of Ljubljana**, Ales Maver^97^, Borut Peterlin^97^

**University of Lübeck**: Alexander Münchau^98^, Katja Lohmann^99^, Rebecca Herzog^98, 100^, Martje Pauly^98, 99^

**Val d’Hebron Barcelona**: Alfons Macaya^101, 102^, Ana Cazurro-Gutiérrez^101^, Belén Pérez-Dueñas^101^, Francina Munell^101^, Clara Franco Jarava^103, 104^, Laura Batlle Masó^105, 106^, Anna Marcé-Grau^101^, Roger Colobran^103, 104, 107^

**Hospital Sant Joan de Déu Barcelona**: Andrés Nascimento Osorio^108^, Daniel Natera de Benito^108^

**University of Freiburg**: Hanns Lochmüller^109-111^, Rachel Thompson^111^, Kiran Polavarapu^111^, Bodo Grimbacher^112-116^

**University of Oxford**: David Beeson^117^, Judith Cossins^117^

**Folkhälsan Research Centre**: Peter Hackman^118^, Mridul Johari^118^, Marco Savarese^118^, Bjarne Udd^118-120^

**University of Cambridge**: Rita Horvath^121^, Patrick F. Chinnery^121, 122^, Thiloka Ratnaike^123^, Fei Gao^121^, Katherine Schon^121, 124^

**Catalan Institute of Oncology, Barcelona**: Gabriel Capella^125^, Laura Valle^125^

**KU Munich**: Elke Holinski-Feder^126^, Andreas Laner^127^, Verena Steinke-Lange^126^

**TU Dresden**: Evelin Schröck^128^, Andreas Rump^128, 129^

**Koç University:** Ayşe Nazlı Başak^130^

**Ghent University Hospital**: Dimitri Hemelsoet^131, 132^, Bart Dermaut^132-134^, Nika Schuermans^132-134^, Bruce Poppe^132-134^, Hannah Verdin^133^

**University Hospital Meyer, Florence**: Davide Mei^135^, Annalisa Vetro^135^, Simona Balestrini^135, 136^, Renzo Guerrini^135^

**KU Leuven**: Kristl Claeys^137, 138^

**LUMC**: Gijs W.E. Santen^139^, Emilia K. Bijlsma^139^, Mariette J.V. Hoffer^139^, Claudia A.L. Ruivenkamp^139^

**Ludwig Boltzmann Institute for Rare and Undiagnosed Diseases, Vienna:** Kaan Boztug^140-144^, Matthias Haimel^140-142^

**Institute of Pathology and Genetics, Gosselies, Belgium**: Isabelle Maystadt^145, 146^

**Technical University Munich**: Isabell Cordts^147^, Marcus Deschauer^147^

**Neurology/Neurogenetics Laboratory University of Crete, Heraklion, Crete, Greece:** Ioannis Zaganas^148^, Evgenia Kokosali^148^, Mathioudakis Lambros^148^, Athanasios Evangeliou^149^, Martha Spilioti^150^, Elisabeth Kapaki^151^, Mara Bourbouli^151^

**IRCCS G. Gaslini**: Pasquale Striano^152, 153^, Federico Zara^153, 154^, Antonella Riva^153, 154^, Michele Iacomino^154, 155^, Paolo Uva^155^, Marcello Scala^152, 153^, Paolo Scudieri^153, 154^

**Cliniques universitaires Saint-Luc (CUSL)**: Maria-Roberta Cilio^156^, Evelina Carpancea^156^, Chantal Depondt^157^, Damien Lederer^158^, Yves Sznajer^159^, Sarah Duerinckx^160^, Sandrine Mary^158^

**Institute of Human Genetics, University Hospital Essen**: Christel Depienne^161, 162^, Andreas Roos^111, 163, 164^

**University of Luxembourg**: Patrick May^165^

### Affiliations

1. Institute of Medical Genetics and Applied Genomics, University of Tübingen, Tübingen, Germany.

2. Centre for Rare Diseases, University of Tübingen, Tübingen, Germany.

3. NGS Competence Center Tübingen (NCCT), University of Tübingen, Tübingen, Germany.

4. Department of Neurodegeneration, Hertie Institute for Clinical Brain Research (HIH), University of Tübingen, Tübingen, Germany.

5. German Center for Neurodegenerative Diseases (DZNE), Tübingen, Germany.

6. Department of Neurology and Epileptology, Hertie Institute for Clinical Brain Research (HIH), University of Tübingen, Tübingen, Germany.

7. Department of Human Genetics, Radboud University Medical Center, Nijmegen, The Netherlands.

8. Donders Institute for Brain, Cognition and Behaviour, Radboud University Medical Center, Nijmegen, The Netherlands.

9. Department of Clinical Genetics, Maastricht University Medical Centre, Maastricht, the Netherlands.

10. Radboud Institute for Molecular Life Sciences, Nijmegen, The Netherlands.

11. Department of Internal Medicine and Radboud Center for Infectious Diseases (RCI), Radboud University Medical Center, Nijmegen, the Netherlands.

12. Center for Molecular and Biomolecular Informatics, Radboud University Medical Center, Nijmegen, the Netherlands.

13. Department of Neurology, Radboud University Medical Center, Nijmegen, The Netherlands.

14. Department of Genetics and Genome Biology, University of Leicester, Leicester, UK.

15. John Walton Muscular Dystrophy Research Centre, Translational and Clinical Research Institute, Newcastle University and Newcastle Hospitals NHS Foundation Trust, Newcastle upon Tyne, UK.

16. Primary Immunodeficiency Group, Translational and Clinical Research Institute, Newcastle University and Newcastle upon Tyne Hospitals NHS Foundation Trust, Newcastle upon Tyne, UK.

17. Division of Evolution, Infection and Genomics, School of Biological Sciences, Faculty of Biology, Medicine and Health, University of Manchester, Manchester M13 9WL, UK.

18. Manchester Centre for Genomic Medicine, St Mary’s Hospital, Manchester University Hospitals NHS Foundation Trust, Health Innovation Manchester, Manchester M13 9WL, UK.

19. Dijon University Hospital, Genetics Department, Dijon, France.

20. Dijon University Hospital, Centre of Reference for Rare Diseases: Development disorders and malformation syndromes, Dijon, France.

21. Inserm - University of Burgundy-Franche Comté, UMR1231 GAD, Dijon, France.

22. Dijon University Hospital, FHU-TRANSLAD, Dijon, France.

23. Dijon University Hospital, GIMI institute, Dijon, France.

24. University of Burgundy-Franche Comté, Dijon Economics Laboratory, Dijon, France.

25. University of Burgundy-Franche Comté, FHU-TRANSLAD, Dijon, France.

26. CNAG-CRG, Centre for Genomic Regulation (CRG), The Barcelona Institute of Science and Technology, Baldiri Reixac 4, Barcelona 08028, Spain.

27. Universitat Pompeu Fabra (UPF), Barcelona, Spain.

28. EURORDIS-Rare Diseases Europe, Sant Antoni Maria Claret 167 - 08025 Barcelona, Spain.

29. EURORDIS-Rare Diseases Europe, Plateforme Maladies Rares, 75014 Paris, France.

30. INSERM, US14 - Orphanet, Plateforme Maladies Rares, 75014 Paris, France.

31. Institut National de la Santé et de la Recherche Medicale (INSERM) U1127, Paris, France.

32. Centre National de la Recherche Scientifique, Unité Mixte de Recherche (UMR) 7225, Paris, France.

33. Unité Mixte de Recherche en Santé 1127, Université Pierre et Marie Curie (Paris 06), Sorbonne Universités, Paris, France.

34. Institut du Cerveau - ICM, Paris, France.

35. Ecole Pratique des Hautes Etudes, Paris Sciences et Lettres Research University, Paris, France.

36. Centre de Référence de Neurogénétique, Hôpital de la Pitié-Salpêtrière, Assistance Publique-Hôpitaux de Paris (AP-HP), Paris, France.

37. Hôpital de la Pitié-Salpêtrière, Assistance Publique-Hôpitaux de Paris (AP-HP), Paris, France.

38. Sorbonne Université, Inserm, Institut de Myologie, Centre de Recherche en Myologie, F-75013 Paris, France.

39. AP-HP, Centre de Référence de Pathologie Neuromusculaire Nord, Est, Ile-de-France, Institut de Myologie, G.H. Pitié-Salpêtrière, F-75013 Paris, France.

40. Institut de Myologie, Equipe Bases de données, G.H. Pitié-Salpêtrière, F-75013 Paris, France.

41. AP-HP, Unité Fonctionnelle de Cardiogénétique et Myogénétique Moléculaire et Cellulaire, G.H. Pitié-Salpêtrière, F-75013 Paris, France.

42. Department of Biology and Medical Genetics, Charles University Prague-2nd Faculty of Medicine and University Hospital Motol, Prague, Czech Republic.

43. Department of Paediatrics and Inherited Metabolic Disorders, First Faculty of Medicine, Charles University and General University Hospital in Prague, Prague, Czech Republic.

44. Department of Ophthalmology, First Faculty of Medicine, Charles University and General University Hospital in Prague, Prague, Czech Republic.

45. Centre for Paediatric Rheumatology and Autoinflammatory Diseases, Department of Paediatrics and Inherited Metabolic Disorders, 1st Faculty of Medicine, Charles University and General University Hospital in Prague, Czech Republic.

46. European Bioinformatics Institute, European Molecular Biology Laboratory, Wellcome Genome Campus, Hinxton, Cambridge, United Kingdom.

47. Jackson Laboratory for Genomic Medicine, Farmington, CT 06032, USA.

48. Florence Nightingale Faculty of Nursing, Midwifery & Palliative Care, King’s College, London, UK.

49. Society and Ethics Research, Connecting Science, Wellcome Genome Campus, Hinxton, UK.

50. Genomics England, Queen Mary University of London, Dawson Hall, EC1M 6BQ, London, UK.

51. MRC Centre for Neuromuscular Diseases and National Hospital for Neurology and Neurosurgery, UCL Queen Square Institute of Neurology, London, UK.

52. Department of Neuromuscular Diseases, UCL Queen Square Institute of Neurology, London, UK.

53. Department of Clinical and Movement Neurosciences, UCL Queen Square Institute of Neurology, University College London, WC1N 3BG.

54. Department of Clinical and Experimental Epilepsy, UCL Queen Square Institute of Neurology, London, UK.

55. Dementia Research Centre, Department of Neurodegenerative Disease, UCL Queen Square Institute of Neurology, London, UK.

56. Dubowitz Neuromuscular Centre, UCL Great Ormond Street Hospital, London, UK.

57. NIHR Great Ormond Street Hospital Biomedical Research Centre, London, United Kingdom.

58. Peripheral Neuropathy Research Group, University of Antwerp, Antwerp, Belgium.

59. Laboratory of Neuromuscular Pathology, Institute Born-Bunge, University of Antwerp, Antwerpen, Belgium.

60. Translational Neurosciences, Faculty of Medicine and Health Sciences, University of Antwerp, Belgium.

61. Neuromuscular Reference Centre, Department of Neurology, Antwerp University Hospital, Antwerpen, Belgium.

62. VIB-CMN, Applied and Translational Neurogenomics Group.

63. Dipartimento di Medicina di Precisione, Università degli Studi della Campania “Luigi Vanvitelli”, Napoli, Italy.

64. Telethon Institute of Genetics and Medicine, Pozzuoli, Italy.

65. Istituto di Scienze Applicate e Sistemi Intelligenti “E.Caianiello” - ISASI - CNR.

66. Unit of Medical Genetics, Department of Medical Sciences, University of Ferrara, Italy.

67. Institute of Human Genetics, Medical Faculty, University of Bonn, Bonn, Germany.

68. Center for Hereditary Tumor Syndromes, University Hospital Bonn, Bonn, Germany.

69. i3S - Instituto de Investigação e Inovação em Saúde, Universidade do Porto, Portugal.

70. IPATIMUP - Institute of Molecular Pathology and Immunology of the University of Porto, Portugal.

71. Faculty of Medicine, University of Porto, Portugal.

72. Doctoral Programme in Biomedicine, Faculty of Medicine, University of Porto, Portugal.

73. Doctoral Programme in BiotechHealth, School of Medicine and Biomedical Sciences, University of Porto, Portugal.

74. Doctoral Programme in Computer Science, Faculty of Sciences, University of Porto, Portugal.

75. CHUSJ, Centro Hospitalar e Universitário de São João, Porto, Portugal.

76. Departament of Genetics, Faculty of Medicine, University of Porto, Portugal.

77. Faculty of Sciences, University of Porto, Portugal.

78. Department of Genetics, Genomics Coordination Center, University Medical Center Groningen, University of Groningen, Groningen, The Netherlands.

79. Department of Genetics, University Medical Center Groningen, University of Groningen, Groningen, The Netherlands.

80. ERN-GENTURIS.

81. ERN-RITA: European Reference Network for Immunodeficiency, Autoinflammatory, Autimmune and Paediatric Rheumatic diseases, Utrecht, Netherlands.

82. Ada Health GmbH, Karl-Liebknecht-Str. 1, 10178 Berlin, Germany.

83. College of Health, Well-being and Life-Sciences, Sheffield Hallam University, Sheffield, UK.

84. Dept of Genetics, Assistance Publique-Hôpitaux de Paris - Université de Paris, Robert DEBRE University Hospital, 48 bd SERURIER, Paris, France.

85. INSERM UMR 1141 “NeuroDiderot”, Hôpital Robert DEBRE, Paris, France.

86. Department of Genetics, Assistance Publique-Hôpitaux de Paris - Sorbonne Université, Pitié-Salpêtrière University Hospital, 83 Boulevard de l’Hôpital, Paris, France.

87. Reference center of rare diseases “intellectuel disability of rare causes”, Paris, France.

88. Institut du Cerveau (ICM), UMR S 1127, Inserm U1127, CNRS UMR 7225, Sorbonne Université, 75013, Paris, France.

89. Univ. Bordeaux, MRGM INSERM U1211, CHU de Bordeaux, Service de Génétique Médicale, F-33000 Bordeaux, France.

90. Laboratoire de Génétique Moléculaire, Service de Génétique Médicale, CHU Bordeaux – Hôpital Pellegrin, Place Amélie Raba Léon, 33076 Bordeaux Cedex, France.

91. Institute of Rare Diseases Research, Spanish Undiagnosed Rare Diseases Cases Program (SpainUDP) & Undiagnosed Diseases Network International (UDNI), Instituto de Salud Carlos III, Madrid, Spain.

92. Molecular Genetics and Functional Genomics, Ospedale Pediatrico Bambino Gesù, IRCCS, Rome, Italy.

93. Med Biotech Hub and Competence Center, Department of Medical Biotechnologies, University of Siena, Italy.

94. Medical Genetics, University of Siena, Italy.

95. Genetica Medica, Azienda Ospedaliero-Universitaria Senese, Italy.

96. Institute of Genomic Medicine and Rare Diseases, Semmelweis University, Budapest, Hungary.

97. Clinical Institute of Genomic Medicine, University Medical Centre Ljubljana, Slovenia.

98. Institute of Systems Motor Science, University of Lübeck, Ratzeburger Allee 160, 23562, Lübeck, Germany.

99. Institute of Neurogenetics, University of Lübeck, Ratzeburger Allee 160, 23562, Lübeck, Germany.

100. Department of Neurology, University Hospital Schleswig Holstein, Ratzeburger Allee 160, 23562, Lübeck, Germany.

101. Pediatric Neurology Research Group, Vall d’Hebron Research Institute, Universitat Autònoma de Barcelona, Barcelona, Spain.

102. Institute of Neuroscience, Universitat Autònoma de Barcelona, Barcelona, Spain.

103. Diagnostic Immunology Research Group, Vall d’Hebron Research Institute (VHIR), Barcelona, Spain.

104. Immunology Division, Genetics Department. Vall d’Hebron University Hospital (HUVH), Barcelona, Spain.

105. Infection in Immunocompromised Pediatric Patients Research Group, Vall d’Hebron Research Institute (VHIR), Barcelona, Spain.

106. Pediatric Infectious Diseases and Immunodeficiencies Unit, Vall d’Hebron University Hospital (HUVH),Barcelona, Spain.

107. Immunology Unit. Department of Cell Biology, Physiology and Immunology. Autonomous University of Barcelona (UAB), Bellaterra, Spain.

108. Neuromuscular Disorders Unit, Department of Pediatric Neurology. Hospital Sant Joan de Déu, Barcelona, Spain

109. Department of Neuropediatrics and Muscle Disorders, Medical Center, Faculty of Medicine, University of Freiburg, Freiburg, Germany.

110. Centro Nacional de Análisis Genómico (CNAG-CRG), Center for Genomic Regulation, Barcelona Institute of Science and Technology (BIST), Barcelona, Spain.

111. Children’s Hospital of Eastern Ontario Research Institute, University of Ottawa, Ottawa, Canada.

112. Institute for Immunodeficiency, Center for Chronic Immunodeficiency (CCI), Medical Center, Faculty of Medicine, Albert-Ludwigs-University of Freiburg, Germany.

113. Clinic of Rheumatology and Clinical Immunology, Center for Chronic Immunodeficiency (CCI), Medical Center, Faculty of Medicine, Albert-Ludwigs-University of Freiburg, Germany.

114. DZIF – German Center for Infection Research, Satellite Center Freiburg, Germany.

115. CIBSS – Centre for Integrative Biological Signalling Studies, Albert-Ludwigs University, Freiburg, Germany.

116. RESIST – Cluster of Excellence 2155 to Hanover Medical School, Satellite Center Freiburg, Germany.

117. Nuffield Department of Clinical Neurosciences, University of Oxford, UK.

118. Folkhälsan Research Centre and Medicum, University of Helsinki, Helsinki, Finland.

119. Tampere Neuromuscular Center, Tampere, Finland.

120. Vasa Central Hospital, Vaasa, Finland.

121. Department of Clinical Neurosciences, University of Cambridge, Cambridge, UK.

122. Medical Research Council Mitochondrial Biology Unit, University of Cambridge, Cambridge, UK.

123. Department of Paediatrics, University of Cambridge, Cambridge, UK.

124. East Anglian Medical Genetics Service, Cambridge University Hospitals NHS Foundation Trust, Cambridge, UK.

125. Bellvitge Biomedical Research Institute (IDIBELL), Barcelona, Spain.

126. Medizinische Klinik und Poliklinik IV – Campus Innenstadt, Klinikum der Universität München, Munich, Germany.

127. MGZ - Medical Genetics Center, Munich, Germany.

128. Institute of Clinical Genetics, University Hospital Carl Gustav Carus, Technical University Dresden, Dresden, Germany.

129. Center for Personalized Oncology, University Hospital Carl Gustav Carus, Technical University Dresden, Dresden, Germany.

130. Koç Universıty,School of Medicine, Translational Medicine Research Center, KUTTAM-NDAL Istanbul Turkey.

131. Dpt. of Neurology, Ghent University Hospital, Ghent, Belgium.

132. Program for Undiagnosed Rare Diseases (UD-PrOZA), Ghent University Hospital, Ghent, Belgium.

133. Center for Medical Genetics, Ghent University Hospital, Ghent, Belgium.

134. Department of Biomolecular Medicine, Faculty of Medicine and Health Sciences, Ghent University, Ghent, Belgium.

135. Neuroscience Department, Children’s Hospital A. Meyer-University of Florence, 50139, Florence, Italy.

136. Department of Clinical and Experimental Epilepsy, UCL Queen Square Institute of Neurology, and Chalfont Centre for Epilepsy, Gerrard Cross, UK.

137. Department of Neurology, University Hospitals Leuven, Leuven, Belgium.

138. Laboratory for Muscle Diseases and Neuropathies, Department of Neurosciences, and Leuven Brain Institute (LBI), KU Leuven - University of Leuven, Leuven, Belgium.

139. Department of Clinical Genetics, Leiden University Medical Center, Leiden, The Netherlands.

140. Ludwig Boltzmann Institute for Rare and Undiagnosed Diseases, Vienna, Austria.

141. St. Anna Children’s Cancer Research Institute (CCRI), Vienna, Austria.

142. CeMM Research Center for Molecular Medicine of the Austrian Academy of Sciences, Vienna, Austria.

143. Department of Pediatrics and Adolescent Medicine, Medical University of Vienna, Vienna, Austria.

144. St. Anna Children’s Hospital, Department of Pediatrics and Adolescent Medicine, Medical University of Vienna, Vienna, Austria.

145. Centre de Génétique Humaine, Institut de Pathologie et de Génétique, Gosselies, Belgium.

146. Département de Médecine, Université de namur (Unamur), Namur, Belgique.

147. Department of Neurology, Klinikum rechts der Isar, Technical University Munich, Munich, Germany.

148. Neurology / Neurogenetics Laboratory University of Crete, Heraklion, Crete, Greece.

149. Aristotle University of Thessaloniki, Thessaloniki, Greece.

150. 1st Department of Neurology, Aristotle University of Thessaloniki, University General Hospital of Thessaloniki, AHEPA, Thessaloniki, Greece.

151. Neurochemistry and Biomarker Unit, 1st Department of Neurology, School of Medicine, National and Kapodistrian University of Athens, Eginition Hospital, Athens, Greece.

152. Pediatric Neurology and Muscular Disease Unit, IRCCS Istituto Giannina Gaslini, Genoa, Italy.

153. Department of Neurosciences, Rehabilitation, Ophthalmology, Genetics, Maternal and Child Health, University of Genoa, Genoa, Italy.

154. Unit of Medical Genetics, IRCCS Istituto Giannina Gaslini, Genoa, Italy.

155. Clinical Bioinformatics, IRCCS Istituto Giannina Gaslini, Genoa, Italy.

156. Pediatric Neurology Department, Saint-Luc University Hospital, Université Catholique de Louvain, Brussels, Belgium.

157. Neurology Department, Erasme Hospital, Université Libre de Bruxelles, Bruxelles, Belgium.

158. Institute of Pathology and Genetics, Charleroi, Belgium.

159. Human Genetics Department, Saint-Luc University Hospital, Université Catholique de Louvain, Brussels, Belgium.

160. Institute of Interdisciplinary Research in Human and Molecular Biology, Human Genetics, IRIBHM, Université Libre de Bruxelles, Brussels, Belgium.

161. Institute of Human Genetics, University Hospital Essen, University Duisburg-Essen, Essen, Germany.

162. Institut du Cerveau et de la Moelle épinière (ICM), Sorbonne Université, UMR S 1127, Inserm U1127, CNRS UMR 7225, F-75013 Paris, France.

163. Department of Pediatric Neurology, Developmental Neurology and Social Pediatrics, Children’s Hospital University of Essen, Essen, Germany.

164. Department of Neurology, Heimer Institute for Muscle Research, University Hospital Bergmannsheil, Ruhr-University Bochum, 44789 Bochum, Germany.

165. Luxembourg Centre for Systems Biomedicine, University of Luxembourg, Esch-sur-Alzette, Luxembourg.

