## Supplementary information 1: Code of Conduct for "A unified data infrastructure to support large-scale rare disease research"

### Solve-RD Code of Conduct

**Version**
**Date**
**History of changes**
**Authors**
**Approved by GA**

#### Definitions

- Investigator: Individual data user affiliated to a Solve-RD beneficiary or associated partner.
- Raw dataset: exome or genome sequencing data, omics data, phenotype data from single individuals which have not been processed through an analysis pipeline.
- Processed dataset: data that are the result of raw data being processed through an analysis pipeline by Solve-RD or Solve-RD partners. It also includes analysis results.
- Solve-RD Sandbox: temporary secure compute environment / private 'cloud' for storage of data as well as tailored and *de novo* bioinformatics analysis in Solve-RD. The Solve-RD sandbox is composed of two parts: 1) an EGA box where raw data is being stored and 2) an European Bioinformatics Institute hosted analysis sandbox which can be used for data analysis. All data in the analysis sandbox (part 2 above) will be deleted on the completion of the Solve-RD project unless Union or Member State law requires storage of the data. Data in the EGA box (part 1 above) will be properly archived for the future at the EGA to enable controlled access distribution.

**Contractual basis**

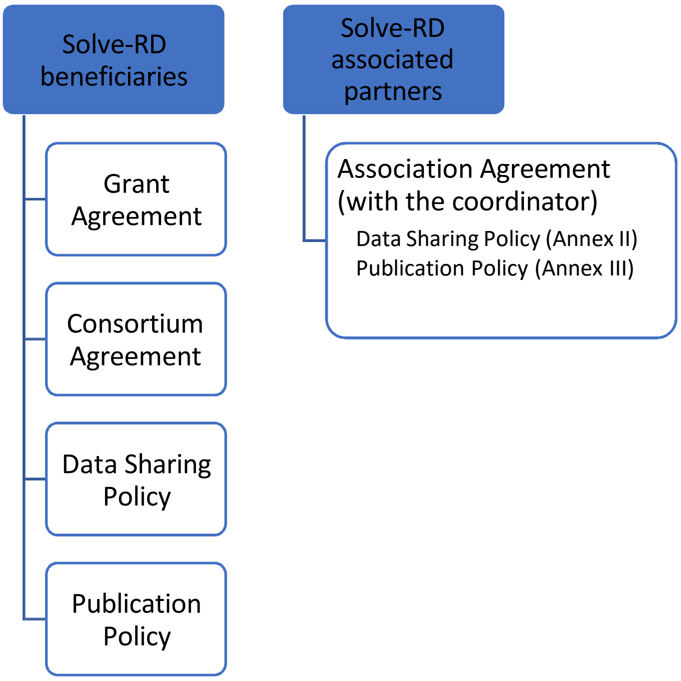


The following is the Code of Conduct that the Investigator agrees to abide by as a user of the Solve-RD analysis sandbox. Failure to abide by any term within this Code of Conduct may result in revocation of approved access to any or all data obtained through the Sandbox.

Solve-RD comprises pseudonymised sensitive data of rare disease patients and their family members (phenotype data, metadata, raw datasets, and processed datasets) that are provided by contributing beneficiaries and associated partners. The contributing beneficiaries and associated partners are the primary data controllers of the datasets they contribute. Data shall not be processed, downloaded and/or used for any analysis unless the specific analysis/usage has been registered through the Solve-RD *Analysis Template* (see attachment 1)* and the Investigator has been granted user rights for the Sandbox. Specifically, Investigators will work according to the following rules:

<u>Principles</u>

1. Investigator will adhere to the Solve-RD Data Sharing Policy[1] (see attachment 2).*
2. Investigator will adhere to the Solve-RD Publication Policy[2] (see attachment 3).*
3. Investigator acknowledges Solve-RD funding in any publication stemming from data processing of any Solve-RD data using the following wording: “The Solve-RD project has received funding from the European Union’s Horizon 2020 research and innovation programme under grant agreement No 779257.”.

<u>Access to the Sandbox</u>

4. Investigator’s permission to access Solve-RD data is limited to all Investigators that have signed this Code of Conduct.

---

1 Latest version available here: http://solve-rd.eu/results/presentations/

2 Latest version available here: http://solve-rd.eu/results/presentations/

* Not added as part of the supplement

5. Investigator will refrain from downloading data to any location that has not been approved by Solve-RD[3].
6. Approvals for downloading data from the Solve-RD Sandbox to secure local clusters will be granted by the Solve-RD Steering Committee upon application from Investigator. Record of all persons locally accessing the data must be kept by the Investigator who has been granted approval and shared with Solve-RD upon request.
7. Downloaded data will have to be deleted on the completion of the Solve-RD project unless Union or Member State law requires storage of the data. Investigator will confirm the deletion of downloaded data and respective processed data to the Solve-RD project office.
8. Investigator will report any inadvertent data release, breach of data security, or other data management incidents contrary to the terms of data access specified in this Code of Conduct to.

Data processing

9. Investigator will perform all analyses solely on the Sandbox, on approved local clusters ensuring data security or on a Solve-RD approved secure space.
10. Investigator will process datasets contained in the Solve-RD Sandbox solely in connection with an approved analysis project described in the Solve-RD Analysis Template (see attachment 1).
11. When data is downloaded from the Sandbox to a local cluster for processing locally, Investigator will ensure that all downloaded data as well as intermediate/output data generated through analysis/data usage, are solely accessible by the Investigator him or herself and their team.
12. Investigator agrees that the data may only be used for the permitted purpose to conduct the approved analysis/data usage.
13. Investigator is not allowed to inform the primary controller of the dataset about an incidental finding, unless the primary controller of that dataset explicitly requests this information from Investigator.
14. Investigator will provide the results of the analysis/data usage to the primary controller of the dataset as soon as the analysis results are available.
15. Investigator is aware that all activities with regards to processing data in the Sandbox and downloading data will be monitored at the level of individual researcher.
16. Investigator will make no attempt to identify or contact participants from whom these data were collected unless re-contact is appropriate for particular reasons. Such decision will be made by the Solve-RD SC. Participants will have the right to withdraw.

**I read and understand the Solve-RD Code of Conduct as well as the Solve-RD Data Sharing Policy and Publication Policy. I declare that I will abide by it and will act accordingly. I understand that failure to adhere to the Code of Conduct will have repercussions for my role in Solve-RD.**

*Please, sign this Code of Conduct and send a scanned copy to the project coordinator*

[3] Download of analysis result files from the DITF folders on the sftp server by DITF and WG members is allowed.
